# Sensorimotor cortex activity during cued gait differs in Parkinson’s freezers and non-freezers

**DOI:** 10.64898/2026.10.19.26365651

**Authors:** Lei Ma, Mai Miura, Ipek Talu, Katy A. Cross

**Author notes:** Correspondence to: Katy A Cross, 710 Westwood Ave, Los Angeles, CA, 90095, USA.

## Abstract

Gait impairments are a major contributor to disability in patients with Parkinson’s disease despite dopaminergic and neuromodulation therapies. External cues (e.g., visual targets, rhythmic auditory sounds) are commonly used to improve gait quality and reduce freezing of gait (FOG), yet their effectiveness varies across patients and contexts. Improved understanding of the neural mechanisms of cueing benefits is essential to personalize rehabilitation strategies and improve outcomes. We use mobile electroencephalography (EEG) to test the hypothesis that cues improve gait through engagement of voluntary motor control mechanisms to compensate for impaired gait automaticity. In addition, we investigate whether this mechanism is related to cueing benefits or freezing status. Eighteen PD patients with a history of freezing (PD-F), 21 patients without freezing (PD-NF) and 19 age-matched healthy controls (HC) walked overground under three conditions: normal walking, synchronizing to auditory cues, and stepping over visual cues. Stride-locked beta band (13–30 Hz) power dynamics in sensorimotor cortex were used to measure cortical control of stepping. Spatiotemporal gait parameters improved with cues in all groups. However, PD-F had smaller cue-related gains in speed and stride length when compared with PD-NF for both cue types, and did not improve gait speed with visual cues. EEG revealed greater contralateral sensorimotor cortex swing-phase beta suppression in PD-F than PD-NF patients, with visual cues eliciting the most robust response. In contrast, beta suppression with cues was less pronounced in PD-NF and HC, despite greater behavioral gains. In addition, engagement of sensorimotor cortex during visual cueing was associated with divergent gait effects across PD subgroups: In PD-NF greater beta suppression was associated with less gait improvement, whereas in PD-F the opposite was observed. These findings suggest distinct sensorimotor control mechanisms for cue utilization in PD-F and PD-NF, and indicate that engagement of voluntary motor control circuits may underlie cueing benefits primarily in PD-F.

## Introduction

Gait impairments are a hallmark of Parkinson’s disease (PD), often presenting as slower gait speed, reduced stride length, and increased stepping variability.^1,2^ Patients may also experience freezing of gait (FOG), a debilitating phenomenon defined as brief, episodic absence or marked reduction of forward progression of the feet despite the intention to walk.^2–4^ These gait disturbances increase fall risk, reduce quality of life, and are often poorly responsive to dopaminergic and neuromodulation therapies.^3,4^ External cues, such as visual stepping targets or rhythmic sounds, are commonly used in rehabilitation to improve gait and alleviate FOG.^5–7^ However, the effectiveness of external cueing varies across individuals with PD and even within the same individual in different contexts.^7,8^ Improved understanding of the neural mechanisms underlying cueing benefits may enable more personalized and effective treatment approaches.^9^ With growing interest in neuromodulation to improve gait function in PD, clarifying cortical modulation during gait could also inform more targeted, evidence-based stimulation parameters to alleviate gait impairments.^10–12^

Normal gait depends on the integration of automatic and voluntary, or goal-directed, processes.^13,14^ Automaticity allows for gait with minimal attentional demands, whereas voluntary control is necessary to adapt gait to task goals or environmental constraints. In PD, degeneration of basal ganglia circuits leads to impairment of automatic movements, forcing patients to rely more heavily on voluntary control of gait.^14,15^ It has been proposed that external cueing improves gait by drawing attention to individual stepping movements, thereby facilitating the engagement of cortical circuits for voluntary movement.^5,16–18^ Consistent with this perspective, neuroimaging studies have demonstrated increased cortical activity during externally cued walking compared to uncued walking in PD.^17–19^ Visual and auditory cues altered activity in sensorimotor, parietal, and occipital regions, with varying patterns across the studies. This work has focused on differences between cued and uncued walking bouts with low temporal resolution (seconds to minutes), leaving precise cortical contributions to stepping execution unexplored. Furthermore, it is unclear if changes in cortical engagement during cued walking reflect adaptive or maladaptive gait control.

Voluntary limb movements, including those during gait, are produced by motor cortex output transmitted via the corticospinal tract.^20–22^ The engagement of motor cortex in movement can be measured non-invasively with scalp EEG as oscillatory neural activity. Oscillatory activity in the beta (13-30Hz) frequency band in the sensorimotor cortex is a robust marker of voluntary motor processes, where movement planning and execution are associated with suppression (or desynchronization) of beta power.^23–25^ Advances in scalp EEG acquisition hardware and signal processing have improved the signal-to-noise ratio of EEG during gait, allowing monitoring of dynamic cortical activity across different phases of the gait cycle.^26,27^ Using this technique, growing evidence demonstrates that the sensorimotor cortex is engaged during gait. Specifically, beta is suppressed during the contralateral swing phase of the gait cycle and functionally coupled with leg muscle activity.^27–33^ Importantly, this beta suppression is augmented when gait demands greater voluntary control of stepping movements, such as walking on uneven surfaces,^34,35^ precision stepping on visual targets,^32^ or synchronizing steps to others.^36^

Here, we test the hypothesis that external cueing enhances gait in PD by increasing voluntary control of stepping, as reflected in beta-band power dynamics over the sensorimotor cortex during gait. We leverage the high temporal resolution of EEG to examine gait-phase-specific beta suppression during two external cueing conditions, visual targets and rhythmic auditory stimuli, to address the possibility that distinct mechanisms may be involved for different types of cues.^9,17^ We predicted that cueing would elicit greater beta suppression over the contralateral sensorimotor cortex during the swing phase, reflecting increased cortical control of stepping. Given that cueing efficacy varies with gait severity and the presence of FOG,^7^ we further investigated whether cue-related beta suppression differs between patients with and without FOG. Finally, we tested whether gait-related beta suppression predicts the magnitude of cueing benefits, hypothesizing that stronger beta suppression would be associated with greater gait improvements due to more successful recruitment of compensatory voluntary motor control.

## Materials and methods

A total of 68 participants (45 PD participants and 23 age-matched healthy controls) were enrolled. Of these, two did not meet inclusion criteria, one withdrew due to fatigue, and seven experienced technical problems during data acquisition. The final sample comprised 21 PD patients without freezing of gait (PD-NF; 5 Female / 16 Male), 18 PD patients with freezing of gait (PD-F; 3 F / 15 M), and 19 healthy controls (HC; 8 F /11 M). Participants with PD performed the experiment in the practically defined “off” state, after overnight withdrawal from dopaminergic medications.

Inclusion criteria for participants with PD were a diagnosis of PD according to the Movement Disorder Society Clinical Diagnostic Criteria,^37^ and age over 18 years. Exclusion criteria included a history of any neurologic diagnosis other than PD, or musculoskeletal or other conditions that impaired walking, use of an assistive device for indoor walking, inability to walk prior to morning PD medications, history of deep brain stimulation (DBS) surgery, dementia, or active hallucinations. Prior to completing the experimental tasks, all participants completed the Montreal Cognitive Assessment (MoCA) to assess global cognitive function,^38^ the Benton Judgement of Line Orientation (even short form) to assess visuospatial function,^39,40^ and the Trail Making Test (TMT) to assess executive function using Part B – Part A to remove the effect of motor speed.^41^ Participants with PD additionally completed the New Freezing of Gait Questionnaire (NFOG-Q)^42^ and the Parkinson Anxiety Scale (PAS),^43^ and a movement disorders specialist administered the motor subsection of the Unified Parkinson’s Disease Rating Scale Part III (UPDRS-III).^44^ Demographic and clinical information is presented in Table 1.

**Table 1.** Demographic Table.

| Variables | HC (n=19) | PD-NF (n=21) | PD-F (n=18) | P-values |
| --- | --- | --- | --- | --- |
| Sex (F:M) | 8:11 | 5:16 | 3:15 | 0.25<sup>f</sup> |
| More Affected Side (L:R) |  | 10:11 | 13:5 | 0.19<sup>f</sup> |
| NFOG-Q |  |  | 9.22 ± 7.07 |  |
| Height | 171.38 ± 9.25 | 172.42 ± 8.71 | 173.93 ± 10.01 | 0.71<sup>f</sup> |
| Weight | 164.21 ± 27.63 | 160.00 ± 30.45 | 168.94 ± 40.92 | 0.71<sup>f</sup> |
| Age | 70.26 ± 7.92 | 73.62 ± 5.92 | 68.39 ± 6.67 | 0.06<sup>f</sup> |
| BENTON | 9.58 ± 3.92 | 10.48 ± 2.48 | 10.56 ± 3.22 | 0.59<sup>f</sup> |
| MOCA | 25.89 ± 2.75 | 24.76 ± 3.91 | 26.17 ± 2.77 | 0.35<sup>f</sup> |
| UPDRS-III |  | 25.19 ± 8.54 | 35.44 ± 12.12 | < 0.01<sup>t</sup> |
| PAS |  | 7.38 ± 5.38 | 12.11 ± 8.43 | 0.04<sup>t</sup> |
| Trails Making Test B-A | 46.89 ± 30.55 | 85.90 ± 80.62 | 84.17 ± 97.76 | 0.2<sup>f</sup> |
| Cadence at baseline (steps/min) | 102.08 ± 9.74 | 100.96 ± 8.8 | 105.49 ± 12.6 | 0.38<sup>f</sup> |
| # of participants in LSM Cluster | 15 | 16 | 15 |  |
| # of participants in RSM Cluster | 13 | 18 | 12 |  |

P-values from ANOVA and Fisher's exact test is denoted as <sup>f</sup>. Independent samples t-test is denoted in <sup>t</sup>. Fisher's exact test was performed for categorical variables: sex and affected side. ± denotes standard deviation.

The study procedures were approved by the UCLA Institutional Review Board (IRB #21-000624). All participants provided informed consent prior to participation and were compensated $25 per hour for their time.

### Walking Task

Participants walked in an outpatient neurology clinic along a designated path under three conditions: normal walking (no-cue), auditory cue, and visual cue (Fig. 1). The total distance was approximately 220 meters in length. For the no-cue condition participants walked at a self-selected comfortable pace. In the auditory cue condition, participants were instructed to step to the beat of metronome tones delivered through wired earphones. The metronome pace was individualized based on each participant’s cadence during the no-cue condition. The patient’s self-selected cadence was used if it was within normative values provided by APDM Moveo Explorer software (103 – 133 steps per minute). If a participant’s baseline cadence was below the normative range (less than 103 steps per minute), the metronome was set 10 percent faster than their baseline. No participant’s cadence was faster than the normative range. This individualized approach ensured that cues were beneficial for gait, since metronome beats that are too slow or too fast may negatively affect gait performance.^45,46^ Before starting the experiment, participants confirmed they could hear the auditory tones, and volumes were adjusted to the patient-reported comfortable level. In the visual cue condition, participants walked while wearing shoe-mounted laser devices (WalkWithPath, Essex, England) attached to both feet.^47^ Each laser projects a line in front of the swinging contralateral limb while the ipsilateral foot is planted. Before starting the experiment, the lasers were adjusted so that participants could comfortably step over the line. Participants confirmed they could see the visual lines and practiced walking with the visual cue until they felt comfortable.

**Figure 1.**
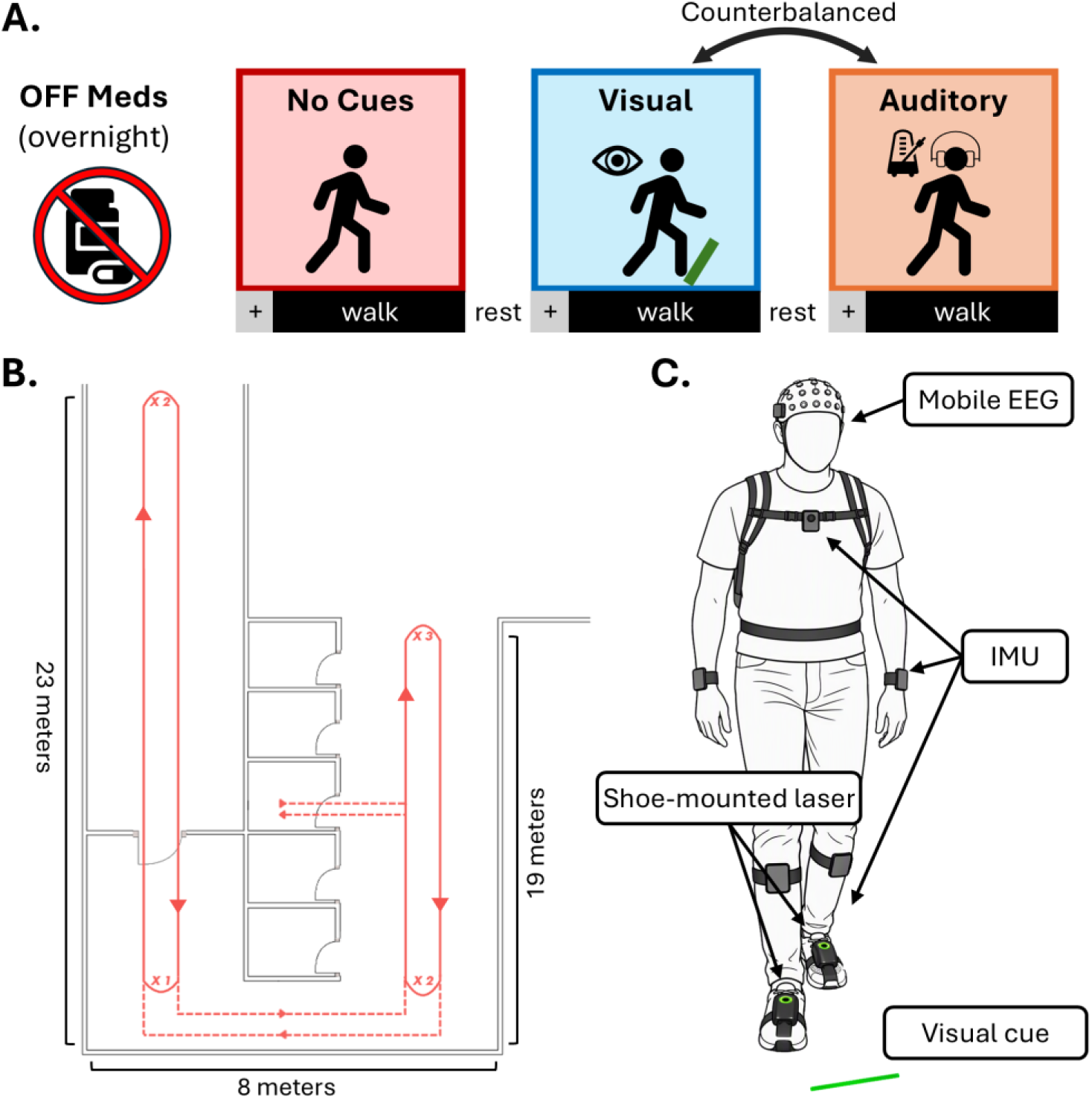
Study Design. (**A**). Study design and task order. Each walking trial began with 30s standing while fixating a cross (+) followed by walking. (**B**) Walking path diagram showing participant route for each walking trial. Not drawn to scale for presentation purposes. Solid lines = multiple passages; dashed lines = single passage. X symbols mark the number of 180-degree turns performed at that location. (**C**) Equipment setup. Eight inertial measurement units (IMU) affixed with Velcro straps, EEG cap and small backpack with EEG amplifier, and shoe-mounted lasers worn in all conditions but activated only during visual cueing.

During the experiment, participants always completed the no-cue condition first to avoid carryover effects of cues. The auditory and visual cue conditions followed, with order counter-balanced across participants. A seated rest break was provided between each condition. Each walking condition began with 30 seconds of quiet standing while participants fixated on a cross at eye level about 5 feet away. The experimenter then verbally cued the participant to walk. The laser device was worn during all conditions but only turned on during the visual cue condition. Each participant performed all three conditions at least once. Conditions were repeated if participants indicated they were not fatigued. The first valid trial for each condition was analyzed; the second attempt was used due to acquisition errors on the first attempt in one PD-F and three PD-NF patients. Data from the visual cue condition is missing for four participants (two HC and two PD-F) due to laser equipment malfunction during the session. Only 3 PD-F experienced FOG episodes during the task and those segments were excluded for current analyses. FOG episodes occurred exclusively during turns and therefore did not affect the quantity of straight-walking data analyzed.

### Spatiotemporal gait parameters

Spatiotemporal gait data were recorded using APDM Opal inertial measurement units (IMUs; APDM Inc., Portland, Oregon), sampled at 128 Hz. Six APDM Opal IMUs were placed on the sternum, fifth lumbar vertebrae, wrists, and dorsum of feet using Velcro straps per manufacturer instructions. Two additional IMUs were placed above the ankles to detect freezing episodes but were not used in analyses due to rare occurrences. Spatiotemporal gait parameters and gait events (i.e., heel strike, toe-off, and turning timestamps) were extracted for each gait cycle using APDM’s Moveo Explorer software.^48,49^ Gait parameters analyzed focused on normalized stride length (cm divided by the height of the participant in cm) and gait speed (cm/s), and the coefficient of variation (%CV) of cadence (steps per minute), representing variability in step timing. These measures were chosen to capture hallmark gait impairments in PD, including reduced stride length, slower gait speed, and increased variability in stepping, that are commonly reported in the PD literature and have been shown to improve with external cueing.^2,9,50^ The APDM system was synchronized with the EEG recordings at the start and end of each walking condition via a digital pulse sent from the APDM system to the EEG amplifier.

### EEG Recording and Preprocessing

Scalp EEG was recorded with a 64-channel equidistant EEG cap connected to the Eego Sports amplifier carried in a small backpack (ANT Neuro, Enschede, Netherlands). Data was sampled at 1024 Hz with impedances kept below 10 kΩ. Offline processing was performed using custom scripts built in Matlab with plugins from EEGLAB,^51^ Fieldtrip,^52^ and others described later in the methods. The pipeline is illustrated in Fig. 2A.

**Figure 2.**
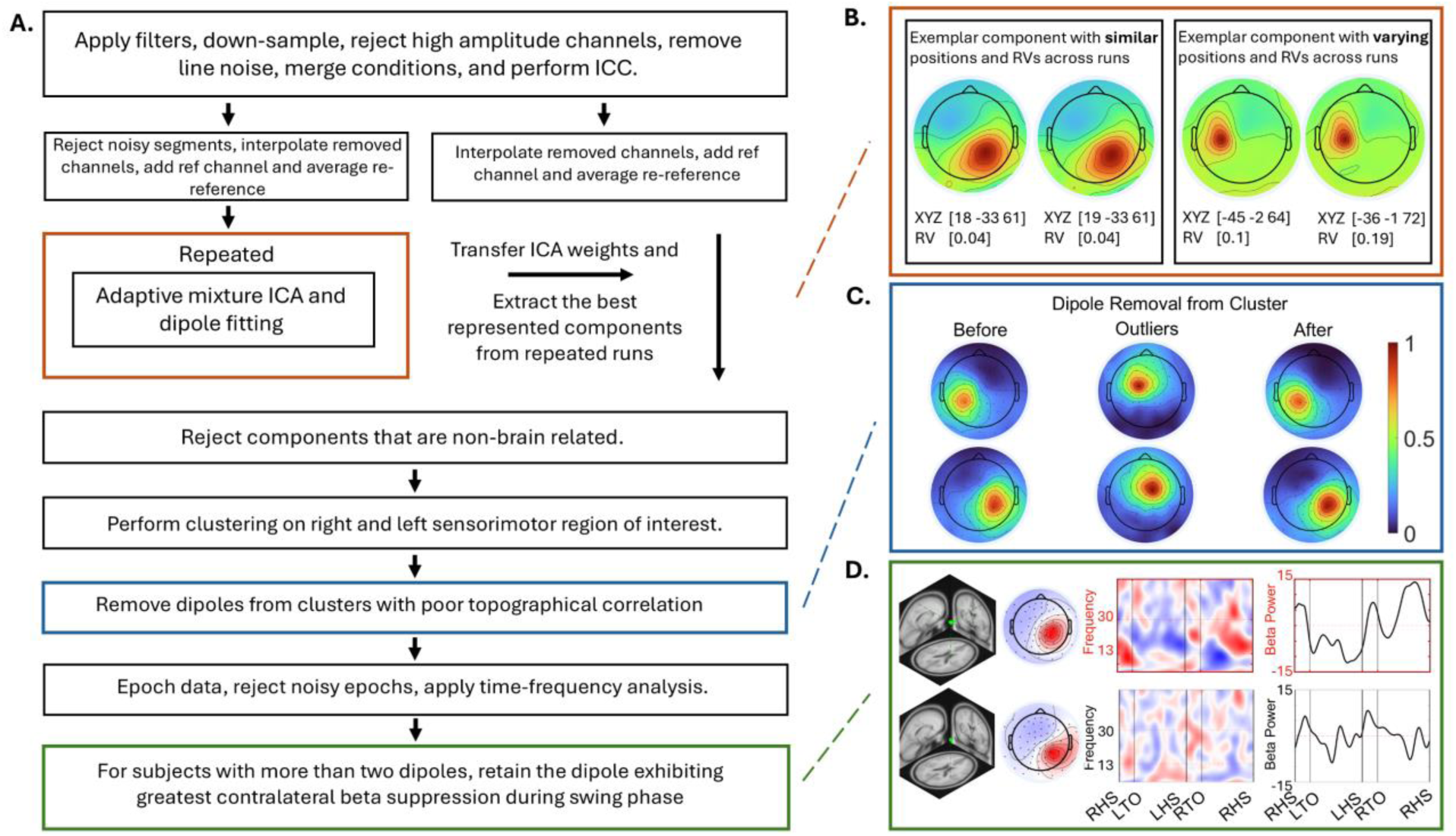
EEG Processing. (**A**) Summary of processing steps to extract left and right sensorimotor dipole sources from raw EEG data. (**B**) Examples of AMICA outcomes for the two dipoles across different iterations. The left example shows a reproduced dipole with similar positions and residual variance (RV) estimates between iterations. The right example shows a reproduced dipole with more varied position and RV estimates between iterations. (**C**) Comparison of the normalized LSM and RSM cluster topographies before and after removal of poorly correlated topographic dipoles. The averaged topography of dipoles with poor correlation is shown in the middle. (**D**) Example of a subject with two dipoles within the same cluster. In such cases, the dipole showing greater beta suppression during the contralateral swing phase averaged across all cycles and conditions was retained for further analysis.

Data were filtered with a low-pass cutoff of 100 Hz (30 Hz transition band) and high-pass cutoff of 1.5 Hz (2 Hz transition band) using zero-phase, non-causal finite-duration impulse response filters, and downsampled to 256 Hz. Zapline-plus plugin was used to remove line noise.^53,54^ A multi-stage process was used to reduce gait-related motion and muscle artifact (Fig. 2). Channels with excessive artifact were identified and removed if the root mean square (RMS) exceeded three standard deviations (SD) from the mean RMS across all the channels in any walking condition.^35,55^ Data were then merged into a single continuous dataset per subject and the iCanClean (ICC) plugin was applied using pseudo-reference signals to clean the data.^26^ These signals are a copy of the original dataset with a temporal band-stop filter (4–50 Hz). Here, ICC treats frequencies below 4 Hz and above 50 Hz as “noise” signals,^26^ and performs canonical correlation analysis to detect and remove any noise components that were highly correlated with these pseudo signals. ICC parameters were set at r-squared > 0.8 with 2 second moving windows. Lastly, EEGLAB’s clean_channels function was applied to remove channels with high-frequency noise (>4 SD), or low correlation with neighboring channels (<0.75). On average, 4.34 (SD = 2.4) channels were removed per participant.

Independent component analysis (ICA) was applied to decompose the EEG data into temporally independent components reflecting neural and non-neural sources.^27,56–59^ A copy of the continuous preprocessed EEG data was created, from which straight-walking segments were extracted based on strides identified by the APDM system. Two complementary cleaning procedures were applied in parallel: EEGLAB’s jointprob function (initial threshold = 3 SD) and clean_windows (window criterion = 0.3; initial tolerance = 5 SD). For each method, thresholds were independently adjusted in 0.5-SD increments until no more than 10% of each participant’s data were flagged from either function. Flagged data segments were removed, previously rejected channels were interpolated, and re-referenced to the average before running adaptive mixture ICA (AMICA).^60^ AMICA was performed using runamica15 function (maximum iterations = 6000), with its sample rejection procedure applied for five iterations, starting after the 2^nd^ iteration.^59^

Source localization on the AMICA results was performed using the EEGLAB’s DIPFIT plugin. Channel locations of each participant were digitized using 3D scans obtained using an iPad Mini (Apple Inc, Cupertino, CA) equipped with a Structure Sensor (Structure, Boulder, CO) during experiment set-up.^61^ The digitized channel locations were co-registered to the standard boundary-element head model in Montreal Neurological Institute (MNI) space for dipole fitting.

ICA solutions can vary slightly across runs, resulting in differences in dipole properties (Fig. 2B) such as location, residual variance (RV), and downstream classification when using heuristic criteria (e.g., using ICLabel or dipole location).^62–64^ Guided by prior solutions to determine ICA reliability through repeated decompositions,^62,63^ AMICA and source localization was repeated 20 times for each participant. ^63^ We compared and clustered independent components (ICs) from all decompositions based on their component activation time series and scalp maps to identify reproducible ICs across AMICA iterations. For each IC pair, we computed Pearson correlation coefficients between the component activations and between mixing matrices (scalp projections) and the smaller of the two was taken as the pair’s similarity to produce a similarity matrix R across all pairs. The distance matrix (1 − R) was submitted to agglomerative average-linkage clustering,^63^ and cluster centrotypes were identified as the IC with the highest summed similarity to cluster members, as previously described.^63^ ICs were assigned to the cluster of their best-matching centrotype, clusters containing ICs from fewer than 19 of the 20 iterations were discarded, and ICs with absolute correlations with the centrotype below r = 0.9 in both measures were flagged as unreliable and excluded from further analyses. On average, 1.2 (SD = 2) clusters per participant were excluded for insufficient consistency across iterations. The activation, weights, dipole location, and RV of each retained cluster’s centrotype were used for further analysis.

To remove non-brain sources, dipoles were deemed artifactual and excluded if they were located outside the brain or had RV greater than 15%;^17,34–36,55,65^ had an upward spectral slope between 5 and 40 Hz suggestive of muscle artifacts;^35^ or exhibited strong cross-frequency power–power coupling in low-frequency (<8 Hz) and high-frequency (>30 Hz) bands, quantified using PowPowCat plugin,^66^ indicative of ocular or muscle activity.^55^ On average, 13.3 (SD = 4.78) dipoles were retained per participant.

#### EEG Spectral Analysis

To extract dynamic gait cycle activity, the cleaned data were epoched from 1 second before to 2 seconds after each right heel strike (RHS) event. Outlier epochs were identified and removed using the jointprob function applied to the dipole activations and the reconstructed channel data from the reminding dipoles, with a threshold set at 3 SD. Spectrograms time-locked to the RHS events (event-related spectral perturbation, ERSP) were calculated using EEGLAB’s newtimef function with Morlet wavelet cycles (2, 0.5), with a pad ratio of 2 between the frequencies of 4 to 50 Hz. To standardize the gait cycle time across cycles and participants, ERSPs were linearly interpolated to the median gait cycle length across all participants to align five key gait events: initial RHS, left toe-off (LTO), left heel strike (LHS), right toe-off (RTO), and the subsequent RHS. For each patient, ERSPs were normalized separately for each condition as the percent change from the mean power across the gait cycle at each frequency during that condition. This condition-specific normalization highlights dynamic power fluctuations across the gait cycle that may be masked by a common baseline.^32^

We also calculated power spectral density during walking relative to standing during each condition for comparison with prior literature.^17,19^ Data were segmented into standing and walking periods. EEGLAB’s spectopo function was applied with 1-second windows and 50% overlap. We first removed the aperiodic (1/f) component from the standing and walking PSDs with the FOOOF toolbox using default parameters.^35,67^ Then the standing-period PSD was subtracted from the walking-period PSD to examine gait-related spectral changes across cueing conditions.^17^

#### Dipole clustering

We employed a priori region-of-interest (ROI) approach to cluster dipoles most likely to reflect gait-related sensorimotor cortical activity for group-level analyses. An anatomical constraint was defined by an ellipsoid centered at [–30, –20, 58] for left sensorimotor (LSM) region and [30, – 20, 58] for right sensorimotor (RSM) based on prior EEG studies.^17,33–35,68^ We used a 30 mm radius in the mediolateral (x) and vertical (z) dimensions, and a narrower 25 mm radius in the anteroposterior (y) direction to minimize overlap with adjacent frontal and parietal regions.^69^ Dipoles within this region whose scalp maps were poorly correlated (r<0.3) with the cluster-level projection (median averaged scalp map across participants) were deemed unlikely to share a similar source and removed (Fig. 2C).^70^ Finally, to prevent artificial sample size inflation, if a participant had more than one dipole in a cluster, we selected the dipole with the greatest total beta suppression during the contralateral relative to ipsilateral swing phase, as this is most likely to represent gait-related sensorimotor cortex activity (Fig. 2D).^27,32,35,36,65^ Circular inference was avoided by pooling all walking conditions in this selection process, making it independent of the contrasts tested between conditions in the statistical models.^71^ This clustering approach mitigates the instability of commonly used K-mean clustering, which can yield variable solutions across runs,^72–74^ by incorporating a priori knowledge into dipole selection.

### Statistical Analysis

#### Spatiotemporal gait parameters

The effects of cue condition and group on gait parameters were investigated with separate linear mixed-effect models for each gait outcome variable (gait speed, stride length, cadence variability). Cue condition (no-cue, auditory, visual), group (HC, PD-NF, PD-F), and their interaction were included as fixed effects, with participant modeled as a random intercept to account for between-subject variability. Age was included as a covariate due to a trend toward baseline group differences (p = 0.06) as well as its strong association with gait decline.^75,76^ If significant cue × group interaction was identified, follow-up interaction contrasts tested whether cue effects (i.e., differences between cue conditions) differed between groups. If there were main effects without an interaction, post-hoc main effect contrasts were conducted for that factor. Additionally, planned *a priori* pairwise contrasts were conducted between cue conditions within each participant group to assess cue-related benefits using estimated marginal means from the models. We interpret uncorrected p-values for these a-priori comparisons but report uncorrected and corrected p-values with FDR across all contrasts within each model for transparency. All post-hoc and pairwise contrasts were two-tailed.

### Sensorimotor Cortex Spectral Power

We first examined the spectral power of the sensorimotor EEG dipole clusters averaged over the full walking bout for each condition, as in previous work.^17,19^ PSD from 4–50 Hz was compared across frequency bins using FieldTrip’s cluster-based permutation tests (2,000 permutations) to control the family-wise error rate across frequency bins. Participants missing any cue condition were excluded (n = 4 per dipole cluster). Main effects of group (collapsed across cue) and cue (collapsed across group) were tested with independent-samples and dependent-samples omnibus F statistics, respectively. The group × cue interaction was tested by computing each participant’s cue-contrast difference spectra (auditory – no cue, visual – no cue, visual – auditory) and comparing them between groups with an omnibus F statistic. Significant omnibus effects were followed by pairwise contrasts using two-sided cluster-based permutation tests.

To confirm the presence of swing phase beta suppression in sensorimotor cortex during the gait cycle,^27,77^ we examined ERSP locked to gait-cycle events, collapsed across groups and cue conditions (all walking trials). Regions that significantly deviated from the mean across the full gait cycle were identified using two-sided cluster correction permutation tests across time and frequency.

To test our primary hypothesis that sensorimotor cortex swing-phase beta-suppression differs by cueing condition and PD phenotype (with and without FOG) we extracted contralateral swing-phase beta power (mean 13–30 Hz power from toe-off to heel-strike of the leg opposite the hemisphere of interest) for each patient and condition. The effect of cue condition and group on swing-phase sensorimotor beta power was evaluated with separate linear mixed models for the left and right hemispheres. As in the models for behavioral data, cue (no-cue, auditory, visual), group (HC, PD-NF, PD-F), and their interaction were included as fixed effects, participant was modeled as a random intercept, and age was included as a covariate. Post-hoc contrasts were performed as described for behavioral models. We also repeated these analyses using only PD groups, this time with separating models by the more- and less-affected hemispheres (rather than left and right hemispheres). This allowed us to account for the asymmetric motor severity characteristic of PD, as well as PD-related motor function and anxiety by including UPDRS III and PAS scores as covariates.

### Brain-Behavior Associations

Lastly, we performed brain-behavior regressions using general linear models to test whether cue-related changes in contralateral swing-phase beta power predicted changes in spatiotemporal gait parameters on the more- and less-affected sides. For each gait parameter, we calculated within-subject changes (cue minus no-cue, i.e. benefit) as dependent measures, and fit separate models with contralateral swing-phase beta power changes, PD subgroup, and their interaction as predictors. Age, UPDRS-III, and PAS were included as covariates. Visual and auditory cues were analyzed in separate models to avoid overparameterization, as the available sample size was insufficient to reliably estimate a three-way beta × cue × group interaction. Significant beta by subgroup interactions were followed up using simple-slope and pairwise slope contrasts, with FDR correction on those contrasts.

Statistical modeling was performed in R. Linear mixed-effects models were built using the *lmerTest* package.^78^ Main and interaction effects from the mixed models were evaluated with Type III analyses of deviance using likelihood-ratio (χ²) tests. To address heteroscedasticity in mixed effects models, the *clubSandwich* package was used to compute cluster-robust variance-covariance estimates with CR2 small-sample correction, providing adjusted standard errors and p-values.^79,80^ Planned contrasts and post hoc comparisons from mixed models used the linear_contrast function from *clubSandwich*, with Satterthwaite’s approximation t-test to account for unequal variances.^79^ General linear models (brain-behavior regressions) were built using the *lmtest* package.^81^ Interaction effects were evaluated with F-test using the *car* package.^82^ Due to heteroscedasticity again these models, the *sandwich* package was used to obtain covariance matrix estimators.^82,83^ Simple slopes and contrasts of slopes were tested with emtrends function from *emmeans* package.^84^ The significance threshold was set at *p* < 0.05 for all statistical testing.

## Results

### Spatiotemporal Gait Parameters

Planned contrasts of cue-related differences within each group are shown in Figure 3(A-C), with corresponding estimates, standard errors, confidence intervals, test statistics, and both corrected and uncorrected p-values provided in Supplementary Table 1. Both visual and auditory cues significantly increased gait speed and stride length relative to no-cue in all groups (*p* < 0.05), except for PD-F where visual cueing did not significantly increase gait speed. Auditory cue additionally reduced cadence variability relative to no-cues in all groups (*p* < 0.05). In contrast, visual cue did not significantly improve cadence variability in any group.

**Figure 3.**
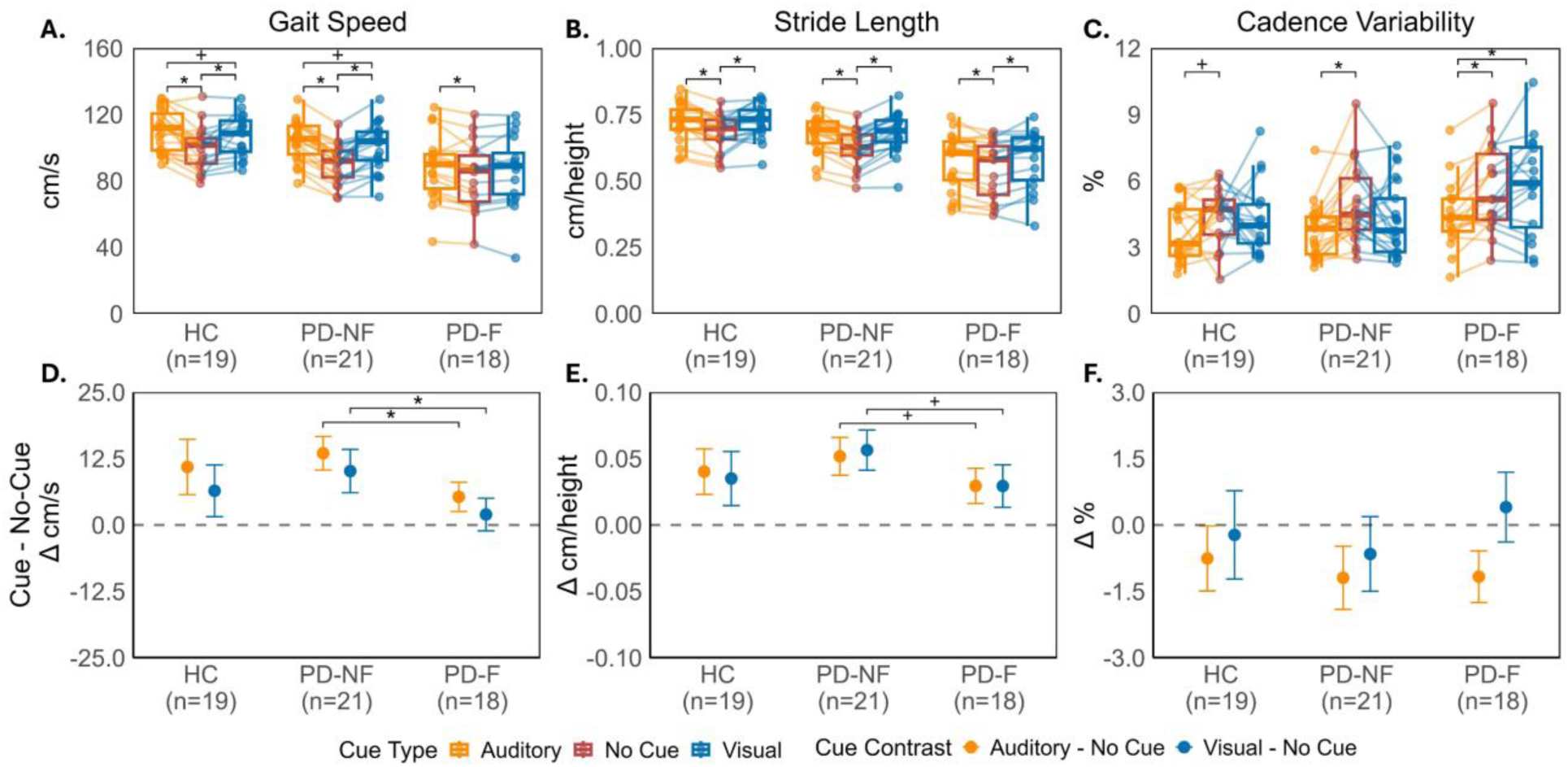
Spatiotemporal gait outcomes. *Top row:* Box and spaghetti plots of gait kinematics for (**A**) gait speed, (**B**) normalized stride length, and (**C**) cadence variability (coefficient of variation %), shown by cue type and group. Lines/dots represent individual subjects. *Bottom row:* Cueing effects shown with estimated mean changes relative to the no-cue condition (i.e., unstandardized effect sizes) for (**D**) gait speed, (**E**) normalized stride length, and (**F**) cadence variability, with error bars representing 95% confidence intervals. Significance bars denote significant contrasts from two-sided Satterthwaite t-test. Statistical significance is indicated by * for FDR-corrected p < 0.05 and + for uncorrected p < 0.05.

There was significant cue by group interaction in the speed model *(χ²(4)* = 24.29, *p* < 0.001). Follow-up interaction contrasts revealed that PD-F benefited less from external cues than PD-NF (Fig. 3D). Specifically, the group difference in cue-related changes showed that PD-F exhibited smaller gains in gait speed than PD-NF (Fig. 3D) with auditory cue compared to no-cue *(difference estimate* = –8.2 cm/s, *95% CI* = [–12.26, –4.15], *t(36.09)* = -4.1*, p* < 0.001*, p.fdr* < 0.01) and with visual cue compared to no-cue *(difference estimate* = –8.17 cm/s, *95% CI* = [–13.11, –3.23], *t(33.86)* = -3.36*, p* < 0.01, *p.fdr* < 0.01).

A significant cue by group interaction was also present in stride length model (*χ²(4)* = 9.59, *p* = 0.048). PD-F showed smaller cue-related gains in normalized stride length compared with PD-NF (Fig. 3E), although these effects were less robust and did not survive correction *(Auditory: difference estimate* = -0.02 cm/height, *95% CI* = [-0.04, 0.00], *t(36.09)* = -2.4, *p* = 0.02*, p.fdr* = 0.1; Visual: *difference estimate* = -0.03 cm/height, *95% CI* = [-0.05, -0.01], *t(33.82) = -2.58 ,p* = 0.01, *p.fdr* = 0.1).

There was no significant interaction for cadence variability, but significant main effects of group (*χ²(2)* = 7.39, *p* = 0.02) and cue (*χ²*(2) = 34.87*, p* < 0.001) were observed. Across cues, cadence variability was lower in HC compared with PD-F *(difference* = -1.35%, *95% CI* = [–2.36, –0.34], *t(33.4) =* -2.72*, p* = 0.01, *p.fdr* = 0.03). Across groups, cadence variability was lower with auditory cue compared to no-cue *(estimate* = -1.04%, *95% CI =* [-1.42, 0.66], t(54.09) = -5.56 , *p* < 0.001, *p.fdr* < 0.001) and compared to visual cue *(estimate* = –0.88%, *95% CI* = [–1.35, –0.42], *t(51.03)* = -3.8, *p* < 0.001, *p.fdr* < 0.001). There were no significant effects of the age covariate in any model.

### Sensorimotor Cortex Spectral Power

PSD and ERSP confirmed expected cortical gait-related activity in the sensorimotor clusters (Fig. 4). There was a consistent reduction in power within and below the beta range during gait relative to standing in the PSD across both clusters, similar to prior reports (Fig. 4B).^17,31^ However, there were no main effects of group, cue or cue by group interactions. ERSP analysis revealed clear lateralized modulation of beta power (13–30 Hz) across the gait cycle, with prominent contralateral beta suppression during the swing phase (Fig. 4C).

**Figure 4.**
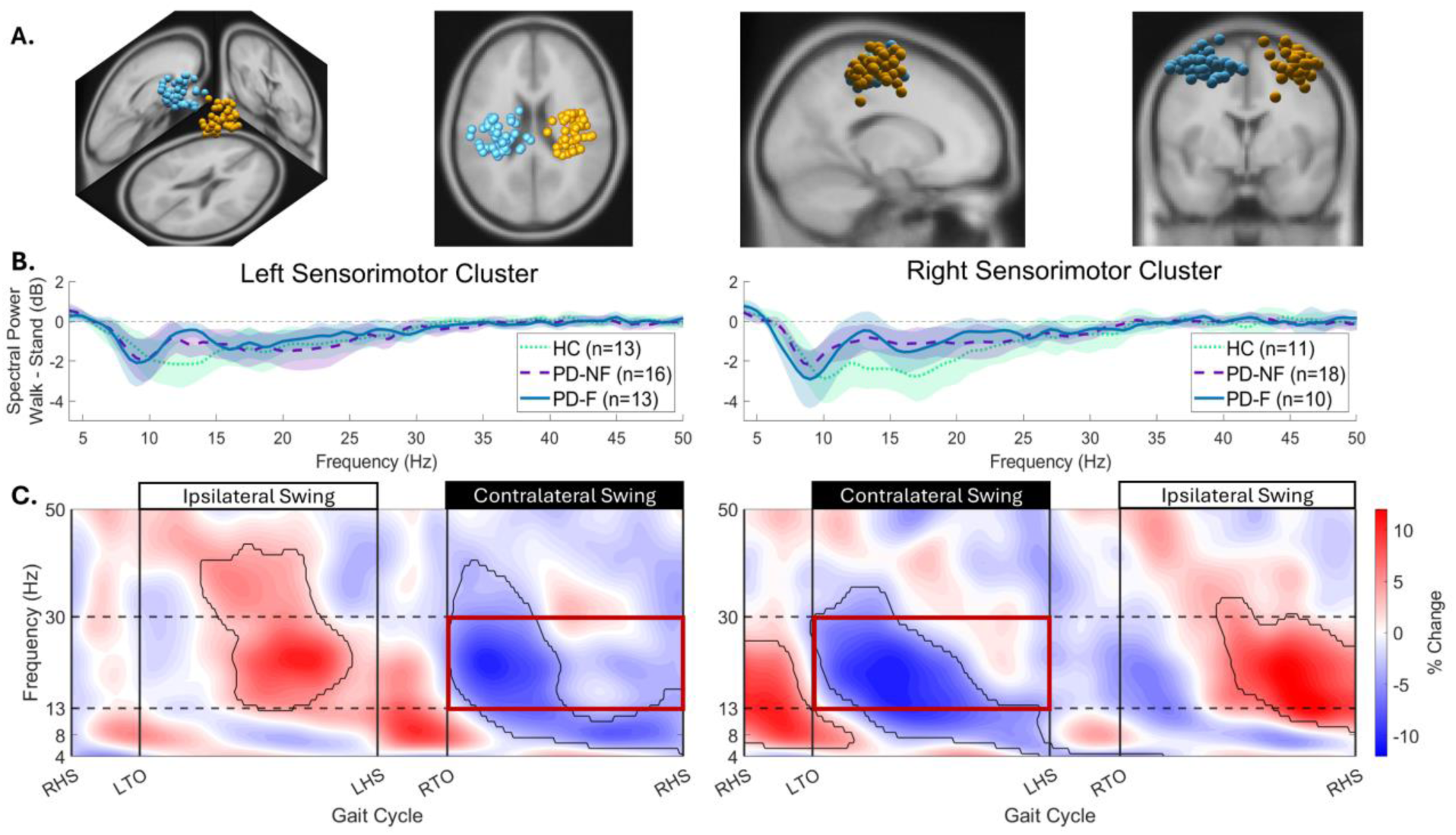
Spectral power in sensorimotor cortex clusters. (**A**) Three-dimensional, axial, sagittal, and coronal views of the left sensorimotor (LSM, blue) and right sensorimotor (RSM, yellow) dipole clusters in template Montreal Neurological Institute (MNI) space. Each dot represents a single dipole localized from an individual participant. (**B**) Power spectral density (PSD) during gait relative to standing for RSM and LSM cluster per group averaged across cue conditions. Shaded areas represent 95% confidence intervals. (**C**) Gait cycle event-related spectral perturbation (ERSP) plots for the LSM and RSM collapsed across all groups and conditions. Black contours denote time-frequency regions significantly deviating from the baseline power (average power across the gait cycle), determined via two-sided cluster-based permutation testing. X-axis ticks represent gait events: right heel strike (RHS), left toe off (LTO), left heel strike (LHS), and right toe off (RTO). In the LSM (n = 46), a significant positive cluster emerged from the mid-point of the left swing phase through to the subsequent RTO (Sum T = 4357.12, p < 0.05) while significant negative clusters were observed during the contralateral (right) swing phase (Sum T = -6197.65, p < 0.05). Conversely, in the RSM (n = 43), a significant positive cluster was identified from the mid-point of the right swing phase through to the subsequent LTO (Sum T = 5721.86, p < 0.05), with negative clusters occurring during the contralateral (left) swing phase (Sum T = -6260.61, p < 0.05). The red outline indicates the specific time-frequency region used to extract contralateral swing-phase beta power for statistical modeling.

The effect of cues on contralateral swing-phase beta power was investigated with planned contrasts comparing the cue conditions within each group (Fig. 5, with corresponding estimates, standard errors, confidence intervals, test statistics, and both corrected and uncorrected p-values reported in Supplementary Table 2). Contralateral beta power during the swing phase was reduced (i.e., greater suppression) during visual cue compared to no-cue in the PD-F group in both hemispheres, and this effect remained in PD-only models accounting for disease severity (*p* < 0.05, Fig. 5A–D). Contralateral swing-phase beta power was also reduced during auditory cue relative to no-cue in the PD-F group, but this effect was restricted to the LSM cortex (*p* < 0.05, Fig. 5A). In the PD-NF group, auditory cue showed reduced contralateral swing-phase beta power relative to no-cues on the less-affected side (*p* < 0.05, Fig. 5D). No significant cue-related effects on contralateral swing-phase beta power were observed in the HC group in any planned contrasts.

**Figure 5.**
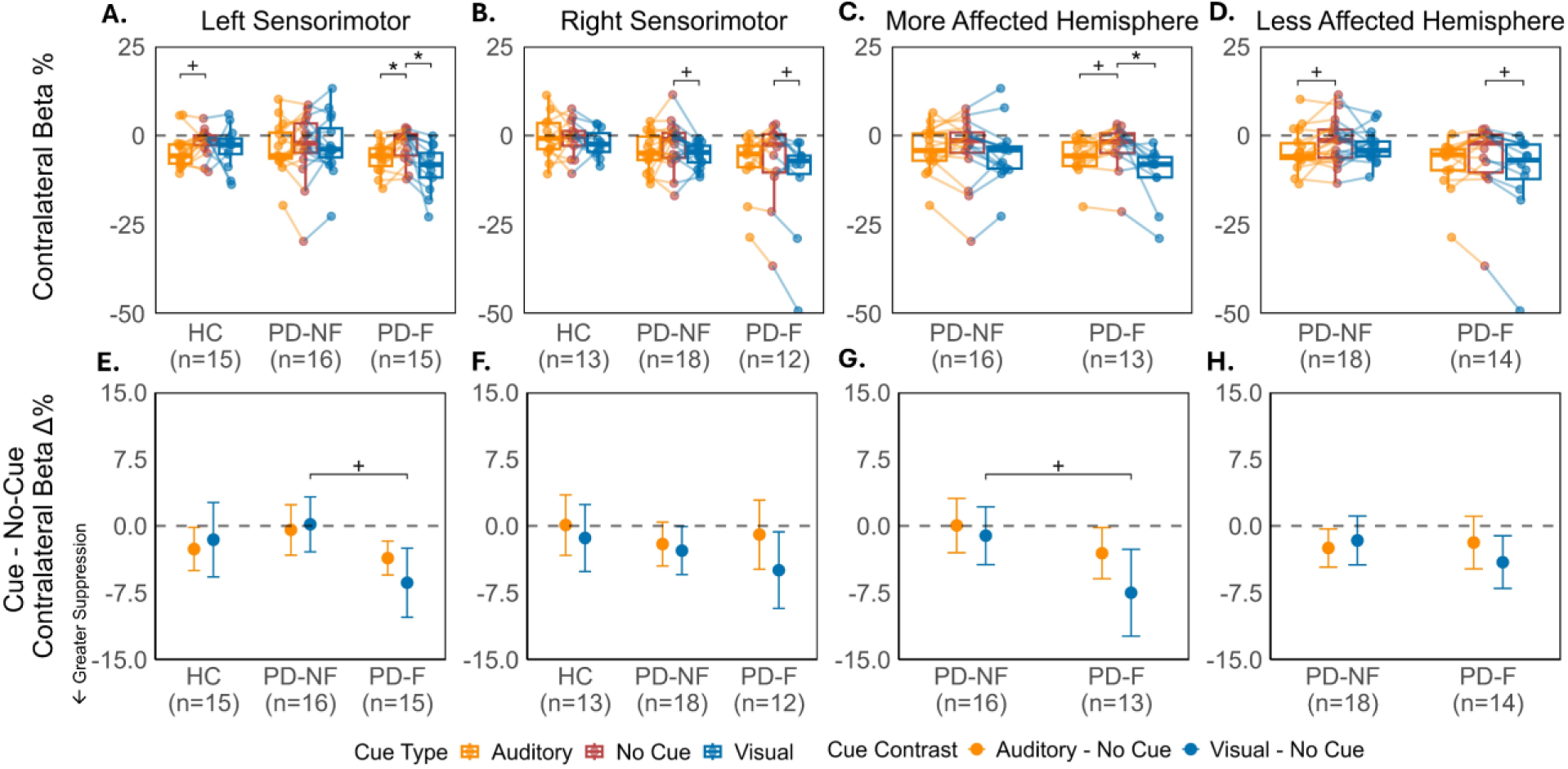
Sensorimotor Contralateral Swing-Phase Beta Power Outcomes. *Top row:* Box and spaghetti plots of contralateral swing-phase beta power (% change from gait cycle average) for (**A**) left sensorimotor, (**B**) right sensorimotor, (**C**) more affected side, and (**D**) less affected side, shown by cue type and group. Lines/dots represent individual subjects. *Bottom row:* Estimated mean changes relative to the no-cue condition (i.e., unstandardized effect sizes) in contralateral swing-phase beta power for (**E**) left sensorimotor, (**F**) right sensorimotor, (**G**) more affected side, and (**H**) less affected side. Significance bars denote significant contrasts from two-sided Satterthwaite t-test. Statistical significance is denoted by * for FDR-corrected p < 0.05 and + for uncorrected p < 0.05.

The RSM and less-affected side models revelated main effects of cue and group on contralateral swing-phase beta power but no interactions. In the RSM, there was a significant main effect of cue, *χ²(2)* = 10.07, *p* < 0.01, with greater beta suppression observed with the visual cue than with no cue (*estimate* = −3.02%, *95% CI* = [−4.97, −1.08], *t(31.26)* = −3.17, *p* < 0.01, *p.fdr* = 0.01). A significant group main effect (*χ²(2*) = 7.08, *p* = 0.03) was due to greater beta suppression in both patient groups relative to HC, but neither difference survived correction (PD-NF: *estimate* = −3.61%, *95% CI* = [−7.16, −0.06], *t(23.98)* = −2.10, *p* = 0.047, *p.fdr* = 0.07; PD-F: *estimate* = −7.20%, *95% CI* = [−13.65, −0.76], *t(21.42)* = −2.32, *p* = 0.03, *p.fdr* = 0.07). In the less-affected side model, there was a significant main effect of cue (*χ²(2)* = 13.14, *p* < 0.01), with greater beta suppression for both the auditory cue and the visual cue relative to no cue (auditory: *estimate* = −2.16%, *95% CI* = [−3.90, −0.43], *t(28.09)* = −2.55, *p* = 0.02, *p.fdr* = 0.02; visual: *estimate* = −2.83%, *95% CI* = [−4.74, −0.92], *t(25.34)* = −3.04, *p* < 0.01, *p.fdr* = 0.02).

A significant cue by group interaction on swing-phase beta power was observed in the LSM model (*χ²(4)* = 10.11, *p* = 0.04). Post-hoc contrasts revealed greater suppression of contralateral swing-phase beta power during visual cue relative to no-cue in PD-F compared to PD-NF, though this did not survive correction for multiple comparisons (*difference estimate* = -6.55%, *95% CI* = [-11.27, -1.82], *t(27.21) =* -2.84*, p* < 0.01, *p.fdr* = 0.08). Similarly, in the PD-only models including clinical covariates, a significant cue × PD subgroup interaction on contralateral swing-phase beta power was observed on the more-affected side model (*χ²*(2) = 6.27, *p* = 0.04). Interaction contrasts revealed that this interaction was driven by greater suppression of contralateral swing-phase beta power during visual cue relative to no-cue in PD-F compared to PD-NF (*difference estimate* = - 6.37%, *95% CI* = [-11.90, -0.85], *t(23.25) = -2.39, p* = 0.03, *p.fdr* = 0.08), but this did not survive correction.

Taken together, the convergent interactions indicate that visual cueing is associated with stronger contralateral swing-phase beta suppression in PD-F than PD-NF patients, particularly in the hemisphere contralateral to the more affected limb. Age was not a significant predictor in any model, however PAS reached significance in the more-affected side model (*χ²(1)* = 5.12, *p* = 0.02).

### Brain-Behavior Associations

Brain–behavior models tested whether greater cue-induced contralateral beta suppression was associated with greater improvements in spatiotemporal gait parameters. On the more affected side during visual cueing, there was a significant subgroup × beta-change interaction in the model predicting gait speed improvements (*F(1,20)* = 8.27, *p* < 0.01), as well as a main effect of subgroup (*F(1,20)* = 7.59, *p* = 0.01). There were no significant main effects of contralateral beta change or covariates (age, UPDRS-III, PAS). Post-hoc slopes analysis showed opposite relationships between beta-change and speed benefits in PD-F and PD-NF groups. Specifically, in PD-NF speed increase was associated with increased swing-phase beta power (less beta suppression) (*estimate* = 0.95 cm/s per percentage-point change in beta, *95% CI* = [0.28, 1.61], *t(20)* = 2.96, *p* < 0.01, *p.fdr* = 0.01), whereas in PD-F the opposite trend was observed *(estimate* = –0.64, *95% CI* = [–1.38, 0.09], *t(20)* = -1.82, *p* = 0.08, *p.fdr* = 0.08) (Fig. 6A). The contrast between the two subgroup slopes remained significant after correction (PD-NF vs. PD-F: *estimate* = 1.59, *95% CI =* [0.44, 2.74], *t(20)* = 2.88, *p* < 0.01, *p.fdr* = 0.01). No other models showed significant relationships between gait improvement and beta change.

**Figure 6.**
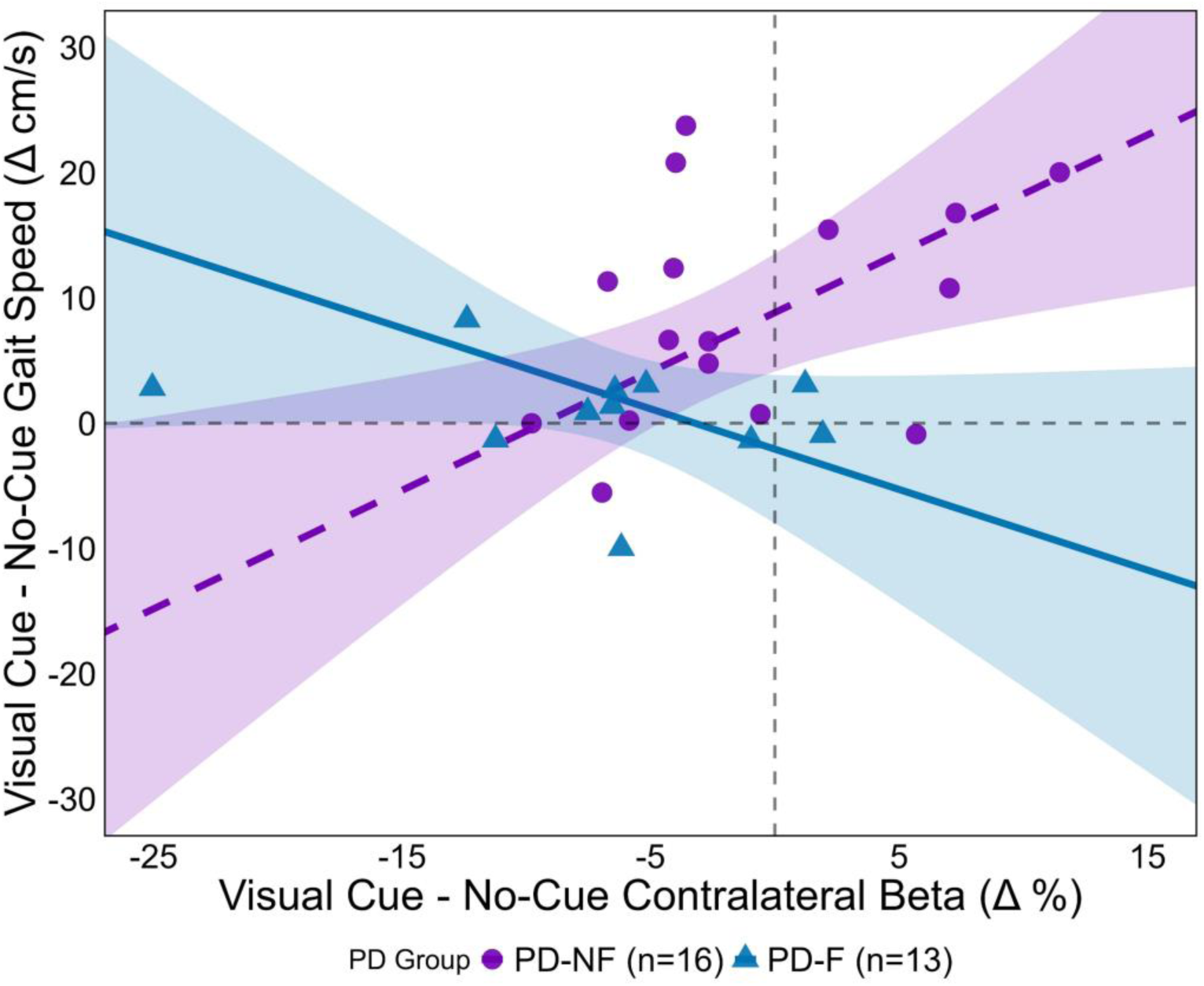
Association between cue-induced changes in brain activity and behavior. Scatterplot illustrates significant relationships between cue-related changes in contralateral sensorimotor swing-phase beta power on the more-affected side and changes in gait speed. Each point represents a single participant’s response to visual cue relative to no-cue. Estimated simple slopes and 95% confidence intervals are shown for PD-F (dark blue, solid line) and PD-NF (purple, dash line). Data points are colored and shaped by group: PD-F (dark blue triangles) and PD-NF (purple circles). Post-hoc pairwise slope contrast revealed a significant difference between PD groups in how contralateral swing-phase beta power relates to cue-related speed improvements. The PD-NF group exhibited a positive correlation between beta power and speed gain, whereas the PD-F group demonstrated an inverse relationship (PD-NF vs. PD-F contrast: estimate = 1.59 cm/s per percentage-point change in beta, 95% CI [0.44, 2.74], t(20) = 2.88, p < 0.01, FDR-corrected p = 0.01). Slopes and the slope contrast were tested with two-sided t-tests. p-values were FDR-corrected across the two group slopes and their contrast.

## Discussion

We investigated whether external cueing promotes voluntary control of stepping, as indexed by beta-band power dynamics over the sensorimotor cortex, and whether this mechanism is related to gait benefits or freezing status in patients with PD. Consistent with previous work, we observed clear engagement of the sensorimotor cortex during gait, characterized by pronounced beta suppression during the contralateral swing phase. Notably, this activity was influenced by external cues and PD subgroups. External cues augmented swing-phase beta suppression, as predicted by the theory that external cueing engages goal-directed gait mechanisms or conscious control of stepping movements.^5,17^ However, this effect was primarily observed in the PD-F group during visual cueing, where cueing benefits were least robust. In contrast, the PD-NF patients and HC showed minimal cue-related swing-phase beta suppression despite greater improvement in stride length and speed than PD-F patients. Thus, while visual cues can engage cortical motor control, this mechanism is not required for improvements in spatiotemporal gait parameters. The contrasting pattern of cue-related beta suppression between PD-F and PD-NF suggests distinct mechanisms for how these groups utilize external cues. Importantly, although PD-F patients exhibited greater motor impairment and higher anxiety than PD-NF patients in the clinical assessments, cue-related contralateral beta differences remained after accounting for these covariates, indicating differences beyond disease severity or anxiety. Instead, these group differences likely reflect distinct strategies for gait adaptation to external cues.

The recruitment of distinct neural mechanisms during externally cued gait in patients with and without freezing of gait may be dictated by the underlying structural and functional integrity of automatic and voluntary gait control networks. Extensive motor learning research shows that shifting attention toward movement goals (an external focus) increases automaticity, reduces reliance on conscious control, and improves motor performance.^85–87^ This framework is especially relevant to PD gait, where automaticity is known to be impaired.^88–90^ External focus has been shown to promote gait automaticity in PD patients.^16,91,92^ However PD-F patients have more severe impairment of gait automaticity than PD-NF patients^89,93^ which may constrain external focus from effectively engaging automatic neural mechanisms in this group. Instead, PD-F patients may be forced to shift to compensatory conscious gait control, which explains the high cortical drive (enhanced beta suppression) and more limited behavioral efficacy of visual cueing in the PD-F group. Conversely, external focus promoted by visual cueing can enhance gait automaticity via the relatively preserved subcortical automatic gait control mechanisms in HC and PD-NF,^94,95^ allowing for efficient utilization of external cues without necessitating increased cortical engagement. Evidence for distinct gait-control mechanisms between PD groups is further suggested by the contrasting relationships between cue-related sensorimotor beta suppression and speed improvement. In PD-NF patients, greater speed gains from visual cues were associated with reduced cortical engagement, consistent with increased automaticity. In contrast, PD-F patients showed greater speed gains with increased cortical engagement, suggesting gait benefits require more conscious control. Together, these findings suggest that visual cueing may promote a shift toward conscious control of stepping when the nervous system’s capacity to engage subcortical automatic mechanisms is impaired.

Despite external cues being routinely employed in PD gait rehabilitation, the neural mechanisms that determine clinical efficacy remain poorly defined. The differences in neural strategies between PD-F and PD-NF groups suggest that tailoring cueing rehabilitation strategies to freezing status may be more efficacious than applying standard cueing protocols. According to our interpretation, in PD-NF patients rehabilitation programs that actively engage and optimize residual gait automaticity may be most effective. In contrast, for PD-F patients, rehabilitation should instead target compensatory cortical motor control pathways. In line with this view, behavioral strategies that promote explicit cortical control through internal focus on movement mechanics also demonstrate superior gait control in patients with freezing compared to externally focused strategies.^96^ Future work should examine whether manipulating attentional focus during cueing produces distinct cortical changes and gait benefits in PD patients or whether neuromodulation approaches targeting compensatory sensorimotor recruitment are more efficacious for PD-F patients.

We included both auditory and visual cues, allowing us also to determine whether the two external cue types improve gait through similar mechanisms. Given partially distinct influences of the two cue types on PD gait parameters (e.g., only auditory cues improved cadence variability), it is likely that auditory and visual cues recruit overlapping but partially distinct neural processes. Here, auditory cueing was associated with reduced contralateral beta power in the LSM of PD-F and the less-affected side of PD-NF, indicating a measurable influence on sensorimotor cortical engagement during PD gait. However, this effect was less robust and spatially inconsistent than the observed visual cueing effects across the hemispheres. One possible explanation is that auditory cueing during gait relies less on visuomotor feedback during the swing phase. This reduced reliance may limit the magnitude of sensorimotor engagement relative to visual cueing in the PD cohorts, which requires continuous online sensorimotor integration to guide stepping toward visual targets. Nonetheless, direct evidence for auditory-driven, gait-phase–specific modulation of neural signals remains limited. Identifying cortical regions that reliably exhibit gait-phase–dependent modulation in response to auditory cues will be essential for achieving a more precise, mechanistic understanding of how external cues differentially shape neural control of stepping.

Our results build on prior cue-related PD EEG studies, which examined PSD changes averaged over several-minute walking bouts under different cueing conditions. Tosserams and colleagues^17^ observed greater beta suppression in response to auditory cues compared to normal walking on a treadmill at constant speed, whereas we did not observe any significant cueing effect on PSD in our sensorimotor clusters. This may be related to differences in treadmill and overground walking, as Vitorio et al^18^ also reported no effect of visual or auditory cues on overground PSD.^18,19^ Treadmill walking at a fixed speed may place different constraints on gait, as changes in cadence are accompanied by corresponding change in stride length, potentially requiring different cortical demand. Nonetheless, Vitorio et al^18^ observed cue-related changes in motor cortical activity using functional near-infrared spectroscopy during visual but not auditory cueing, supporting sensorimotor interaction to visual cues. The absence of sensorimotor EEG changes in prior works may partly reflect the use of PSD measures that collapse neural activity across entire walking trials. Such averaging can obscure transient, phase-specific dynamics that may be associated with discrete stepping events and cue-related modulations. Despite this, Stuart et al^19^ reported early evidence of beta suppression in PSD response to visual cues in patients with PD, primarily localized to centrally clustered EEG dipoles. Methodological differences in their channel density and analytic focus may have limited their spatial specificity of their EEG sources,^56,97^ as well as revealing gait phase-specific dynamics. By leveraging the higher signal-to-noise ratio of high-density EEG, we extracted gait-phase-locked beta dynamics within sensorimotor regions. The targeted gait-cycle–level analyses are better suited to capturing transient neural dynamics associated with individual stepping movements,^27^ and reveal novel insights that extends prior evidence on the cortical mechanisms supporting PD gait.

Several methodological limitations should be acknowledged. The PD patients performed the experiment after overnight withdraw from their regular dopaminergic medications in the clinically “Off” state. This may have amplified their reliance on external cues, possibly explaining why the current study found benefits from shoe-laser cues whereas recent work reported no improvement in PD patients during their “On” medication state.^98^ Future studies will be required to determine whether compensatory mechanisms differ on medications. The APDM Opal IMUs use proprietary algorithms that limit transparency of spatiotemporal metric derivation and IMUs have known limitations in position estimations due to drift. However, prior validation of these metrics in PD and other patient populations suggest adequate measurement properties.^99,100^ Mobile EEG recordings obtained during naturalistic walking are inherently susceptible to motion and muscle artifacts. To mitigate these influences, we implemented rigorous preprocessing steps and focused analyses on the beta frequency band, a well-established marker of motor control in PD. Additionally, group-level dipole clustering presents known challenges. Both inter- and intra-individual variability in dipole localization and clustering parameters can influence cluster quality and compromise localization. Therefore, our clustering and interpretations were intentionally constrained to functional regions of sensorimotor cortex, where our a-priori hypotheses were informed by previously reported robust gait-related effects. As methods for group-level source analysis in mobile EEG continue to evolve, there remains a need for more robust approaches to reliability identify behaviorally relevant dipole clusters.

Despite these limitations, our findings demonstrate that mobile EEG can reliably capture neural dynamics of gait control under ecologically valid walking conditions. Our findings offer the first direct evidence that external cues augment cortical motor control differently in PD patients with and without FOG during the swing phase of the gait cycle. While PD-NF patients achieved gait improvements with limited sensorimotor recruitment, PD-F patients may require increased sensorimotor engagement to obtain gait benefits. Clinically, these insights indicate that rehabilitation for PD-F patients could benefit from strategies that explicitly target and optimize this compensatory cortical control, rather than applying standard cueing protocols designed to restore automaticity. Moving from uniform interventions to approaches guided by individual neural compensation patterns may ultimately yield more consistent functional outcomes. Future studies should therefore prioritize mechanistically driven rehabilitation designs, ensuring that gait therapies are neuro-physiologically targeted.

## Data Availability

The data that support the findings of this study are available from the corresponding author, upon reasonable request.

## Data availability

The data that support the findings of this study are available from the corresponding author, upon reasonable request. Analysis code is available at the following GitHub repository: https://github.com/Cross-Lab-UCLA/Sensorimotor-EEG-responses-to-cueing-in-PD

## Acknowledgements

We would like to thank all research participants who dedicated time and energy to making this study possible. We also thank Maria Paulino, Akash Rathi, Konmal Ali, and Federico Gennaro for assistance with data acquisition.

