## Supplementary Tables for "Sensorimotor cortex activity during cued gait differs in Parkinson’s freezers and non-freezers"

### for

**Supplementary Table 1. Cue Contrasts on Spatiotemporal Gait Parameters.** **2**

**Supplementary Table 2. Cue Contrasts on Contralateral Swing Beta.** **3**

### Supplementary Table 1. Cue Contrasts on Spatiotemporal Gait Parameters.

| Variable | Contrast | Subgroup | Est | SE | CI95_L | CI95_U | t | df | p | p.fdr |
|---|---|---|---|---|---|---|---|---|---|---|
| Gait Speed (cm/s) | Auditory - No Cue | HC | 10.906 | 2.474 | 5.709 | 16.104 | 4.409 | 18 | **0** | **0.001** |
| | Visual - No Cue | HC | 6.423 | 2.309 | 1.548 | 11.298 | 2.782 | 16.804 | **0.013** | **0.023** |
| | Auditory - Visual | HC | 4.483 | 2.096 | 0.057 | 8.909 | 2.139 | 16.804 | **0.047** | 0.061 |
| | Auditory - No Cue | PD-NF | 13.491 | 1.515 | 10.331 | 16.65 | 8.907 | 20 | **0** | **0** |
| | Visual - No Cue | PD-NF | 10.14 | 1.955 | 6.062 | 14.218 | 5.187 | 20 | **0** | **0** |
| | Auditory - Visual | PD-NF | 3.351 | 1.483 | 0.257 | 6.444 | 2.26 | 20 | **0.035** | 0.053 |
| | Auditory - No Cue | PD-F | 5.286 | 1.306 | 2.532 | 8.041 | 4.049 | 17 | **0.001** | **0.002** |
| | Visual - No Cue | PD-F | 1.966 | 1.442 | -1.094 | 5.026 | 1.363 | 15.801 | 0.192 | 0.192 |
| | Auditory - Visual | PD-F | 3.32 | 2.226 | -1.404 | 8.044 | 1.492 | 15.801 | 0.156 | 0.175 |
| Stride Length (cm/height) | Auditory - No Cue | HC | 0.04 | 0.008 | 0.023 | 0.057 | 4.918 | 18 | **0** | **0** |
| | Visual - No Cue | HC | 0.035 | 0.01 | 0.015 | 0.055 | 3.622 | 16.783 | **0.002** | **0.003** |
| | Auditory - Visual | HC | 0.005 | 0.01 | -0.015 | 0.025 | 0.538 | 16.783 | 0.597 | 0.672 |
| | Auditory - No Cue | PD-NF | 0.052 | 0.007 | 0.037 | 0.066 | 7.559 | 20 | **0** | **0** |
| | Visual - No Cue | PD-NF | 0.056 | 0.007 | 0.041 | 0.072 | 7.762 | 20 | **0** | **0** |
| | Auditory - Visual | PD-NF | -0.005 | 0.006 | -0.017 | 0.008 | -0.796 | 20 | 0.436 | 0.56 |
| | Auditory - No Cue | PD-F | 0.029 | 0.006 | 0.016 | 0.043 | 4.68 | 17 | **0** | **0** |
| | Visual - No Cue | PD-F | 0.029 | 0.007 | 0.013 | 0.045 | 3.888 | 15.778 | **0.001** | **0.002** |
| | Auditory - Visual | PD-F | 0 | 0.009 | -0.02 | 0.02 | 0.008 | 15.778 | 0.993 | 0.993 |
| Cadence Variability (%) | Auditory - No Cue | HC | -0.757 | 0.35 | -1.491 | -0.022 | -2.164 | 18 | **0.044** | 0.099 |
| | Visual - No Cue | HC | -0.225 | 0.472 | -1.222 | 0.772 | -0.476 | 16.961 | 0.64 | 0.64 |
| | Auditory - Visual | HC | -0.532 | 0.451 | -1.483 | 0.419 | -1.18 | 16.961 | 0.254 | 0.327 |
| | Auditory - No Cue | PD-NF | -1.194 | 0.342 | -1.907 | -0.481 | -3.494 | 20 | **0.002** | **0.007** |
| | Visual - No Cue | PD-NF | -0.656 | 0.404 | -1.498 | 0.186 | -1.626 | 20 | 0.12 | 0.179 |
| | Auditory - Visual | PD-NF | -0.538 | 0.316 | -1.197 | 0.121 | -1.703 | 20 | 0.104 | 0.179 |
| | Auditory - No Cue | PD-F | -1.169 | 0.276 | -1.752 | -0.587 | -4.236 | 17 | **0.001** | **0.005** |
| | Visual - No Cue | PD-F | 0.402 | 0.372 | -0.387 | 1.192 | 1.081 | 15.968 | 0.296 | 0.332 |
| | Auditory - Visual | PD-F | -1.572 | 0.423 | -2.468 | -0.676 | -3.719 | 15.968 | **0.002** | **0.007** |

**Table S1. Pairwise cue contrasts within each subgroup by spatiotemporal gait parameters.** Estimates (Est) are differences between estimated marginal means, expressed in the units of each measure, from separate mixed-effects models of gait speed, stride length, and cadence variability. Contrasts were tested with robust Wald t-tests (t = Est/SE) using CR2 standard errors (SE) clustered on participant with Satterthwaite degrees of freedom (df); 95% confidence intervals (CI95) derived from the same estimator. p.fdr computed from Benjamini–Hochberg correction across the nine contrasts (3 cue pairs × 3 subgroups) within each Variable. HC = healthy controls; PD-NF = PD without freezing of gait; PD-F = PD with freezing of gait; L/U = lower/upper confidence limits.

**Supplementary Table 2. Cue Contrasts on Contralateral Swing Beta.**

| Variable | Contrast | Subgroup | Est | SE | CI95_L | CI95_U | t | df | p | p.fdr |
|---|---|---|---|---|---|---|---|---|---|---|
| LSM Contralateral Swing Beta (%) | Auditory - No Cue | HC | -2.585 | 1.127 | -5.001 | -0.169 | -2.295 | 14.000 | **0.038** | 0.113 |
| | Visual - No Cue | HC | -1.540 | 1.928 | -5.709 | 2.629 | -0.799 | 12.884 | 0.439 | 0.790 |
| | Auditory - Visual | HC | -1.045 | 1.615 | -4.537 | 2.446 | -0.647 | 12.884 | 0.529 | 0.793 |
| | Auditory - No Cue | PD-NF | -0.446 | 1.322 | -3.263 | 2.371 | -0.337 | 15.000 | 0.741 | 0.833 |
| | Visual - No Cue | PD-NF | 0.180 | 1.441 | -2.892 | 3.252 | 0.125 | 15.000 | 0.902 | 0.902 |
| | Auditory - Visual | PD-NF | -0.626 | 1.556 | -3.942 | 2.691 | -0.402 | 15.000 | 0.693 | 0.833 |
| | Auditory - No Cue | PD-F | -3.601 | 0.882 | -5.492 | -1.710 | -4.084 | 14.000 | **0.001** | **0.010** |
| | Visual - No Cue | PD-F | -6.367 | 1.796 | -10.251 | -2.482 | -3.544 | 12.892 | **0.004** | **0.016** |
| | Auditory - Visual | PD-F | 2.766 | 1.891 | -1.324 | 6.855 | 1.462 | 12.892 | 0.168 | 0.377 |
| RSM Contralateral Swing Beta (%) | Auditory - No Cue | HC | 0.101 | 1.553 | -3.284 | 3.485 | 0.065 | 12.000 | 0.949 | 0.949 |
| | Visual - No Cue | HC | -1.354 | 1.700 | -5.103 | 2.396 | -0.796 | 10.806 | 0.443 | 0.665 |
| | Auditory - Visual | HC | 1.454 | 1.831 | -2.584 | 5.493 | 0.795 | 10.806 | 0.444 | 0.665 |
| | Auditory - No Cue | PD-NF | -2.030 | 1.163 | -4.484 | 0.424 | -1.746 | 17.000 | 0.099 | 0.242 |
| | Visual - No Cue | PD-NF | -2.759 | 1.275 | -5.448 | -0.069 | -2.164 | 17.000 | **0.045** | 0.202 |
| | Auditory - Visual | PD-NF | 0.729 | 1.253 | -1.914 | 3.371 | 0.582 | 17.000 | 0.568 | 0.665 |
| | Auditory - No Cue | PD-F | -0.973 | 1.760 | -4.847 | 2.901 | -0.553 | 11.000 | 0.591 | 0.665 |
| | Visual - No Cue | PD-F | -4.959 | 1.919 | -9.247 | -0.671 | -2.584 | 9.797 | **0.028** | 0.202 |
| | Auditory - Visual | PD-F | 3.986 | 2.249 | -1.038 | 9.010 | 1.773 | 9.797 | 0.107 | 0.242 |
| Less Affected Contralateral Beta (%) | Auditory - No Cue | PD-NF | -2.471 | 1.01 | -4.602 | -0.341 | -2.447 | 17 | **0.026** | 0.077 |
| | Visual - No Cue | PD-NF | -1.624 | 1.295 | -4.356 | 1.107 | -1.255 | 17 | 0.227 | 0.326 |
| | Auditory - Visual | PD-NF | -0.847 | 1.216 | -3.413 | 1.719 | -0.697 | 17 | 0.496 | 0.496 |
| | Auditory - No Cue | PD-F | -1.858 | 1.365 | -4.806 | 1.091 | -1.361 | 13 | 0.197 | 0.326 |
| | Visual - No Cue | PD-F | -4.036 | 1.331 | -6.943 | -1.129 | -3.031 | 11.777 | **0.011** | 0.064 |
| | Auditory - Visual | PD-F | 2.179 | 1.887 | -1.942 | 6.3 | 1.154 | 11.777 | 0.271 | 0.326 |
| More Affected Contralateral Beta (%) | Auditory - No Cue | PD-NF | 0.051 | 1.426 | -2.989 | 3.091 | 0.036 | 15 | 0.972 | 0.972 |
| | Visual - No Cue | PD-NF | -1.096 | 1.519 | -4.334 | 2.141 | -0.722 | 15 | 0.482 | 0.578 |
| | Auditory - Visual | PD-NF | 1.147 | 1.568 | -2.196 | 4.49 | 0.732 | 15 | 0.476 | 0.578 |
| | Auditory - No Cue | PD-F | -3.053 | 1.314 | -5.917 | -0.189 | -2.322 | 12 | 0.039 | 0.116 |
| | Visual - No Cue | PD-F | -7.468 | 2.209 | -12.341 | -2.596 | -3.381 | 10.799 | **0.006** | **0.038** |
| | Auditory - Visual | PD-F | 4.416 | 2.125 | -0.271 | 9.103 | 2.078 | 10.799 | 0.062 | 0.125 |

**Table S2. Pairwise cue contrasts within each subgroup by contralateral swing beta of each hemisphere.** Estimates (Est) are differences between estimated marginal means from mixed-effects models of contralateral swing beta power fitted separately per hemisphere (i.e., Variable). Contrasts were tested with robust Wald t-tests (t = Est/SE) using CR2 standard errors (SE) clustered on participant with Satterthwaite degrees of freedom (df); 95% confidence intervals (CI95) derived from the same estimator. p.fdr computed on Benjamini–Hochberg correction across the contrasts within each Variable. HC = healthy controls; PD-NF = PD without freezing of gait; PD-F = PD with freezing of gait. L/U = lower/upper confidence limits.
